# High density EEG and deep learning improves outcome prediction on the first day of coma after cardiac arrest

**DOI:** 10.1101/2025.01.14.25320516

**Authors:** Andria Pelentritou, Lucas Gruaz, Manuela Iten, Matthias Haenggi, Frederic Zubler, Andrea O Rossetti, Marzia De Lucia

## Abstract

We assessed outcome prediction of comatose patients using a deep learning analysis applied to resting EEG on the first and second day after cardiac arrest (CA), and its added value to clinical prognosis. We recorded 62-channel resting-state EEG in comatose patients after CA across three Swiss hospitals during the first (N=165) and second (N=100) coma day. Patient outcome was classified as favorable if the best Cerebral Performance Category was 1-2. A convolutional neural network provided a predicted probability for favorable outcome for each patient’s and recording day’s 62-channel and 19-channel EEG. Predictive performance was additionally evaluated on an external 19-channel dataset collected outside Switzerland (N=60). The deep learning prediction was compared to EEG-based clinical markers - according to the American Clinical Neurophysiology Society -, brainstem reflexes and motor responses. On the first day, patient outcome was predicted with an accuracy of 0.94±0.03 for 62 channels and 0.90±0.03 and 0.87 for 19 channels using the Swiss and external dataset, respectively. High outcome prediction (0.98 accuracy) was observed when considering only patients with uncertain prognosis based on clinical assessment. The second day was less predictive, with an accuracy of 0.72±0.05. The estimated outcome prediction correlated with spectral power on the first day for favorable (r =0.38, p=0.01) and unfavorable (r=-0.28, p=0.02) outcome patients, and was consistent with clinical markers (p<0.0001), except brainstem reflexes. On the first day of coma in CA patients, a deep learning analysis of resting-state high-density EEG provides accurate outcome prediction, superior to lower-density EEG, and complements clinical markers.

**Graphical abstract:** 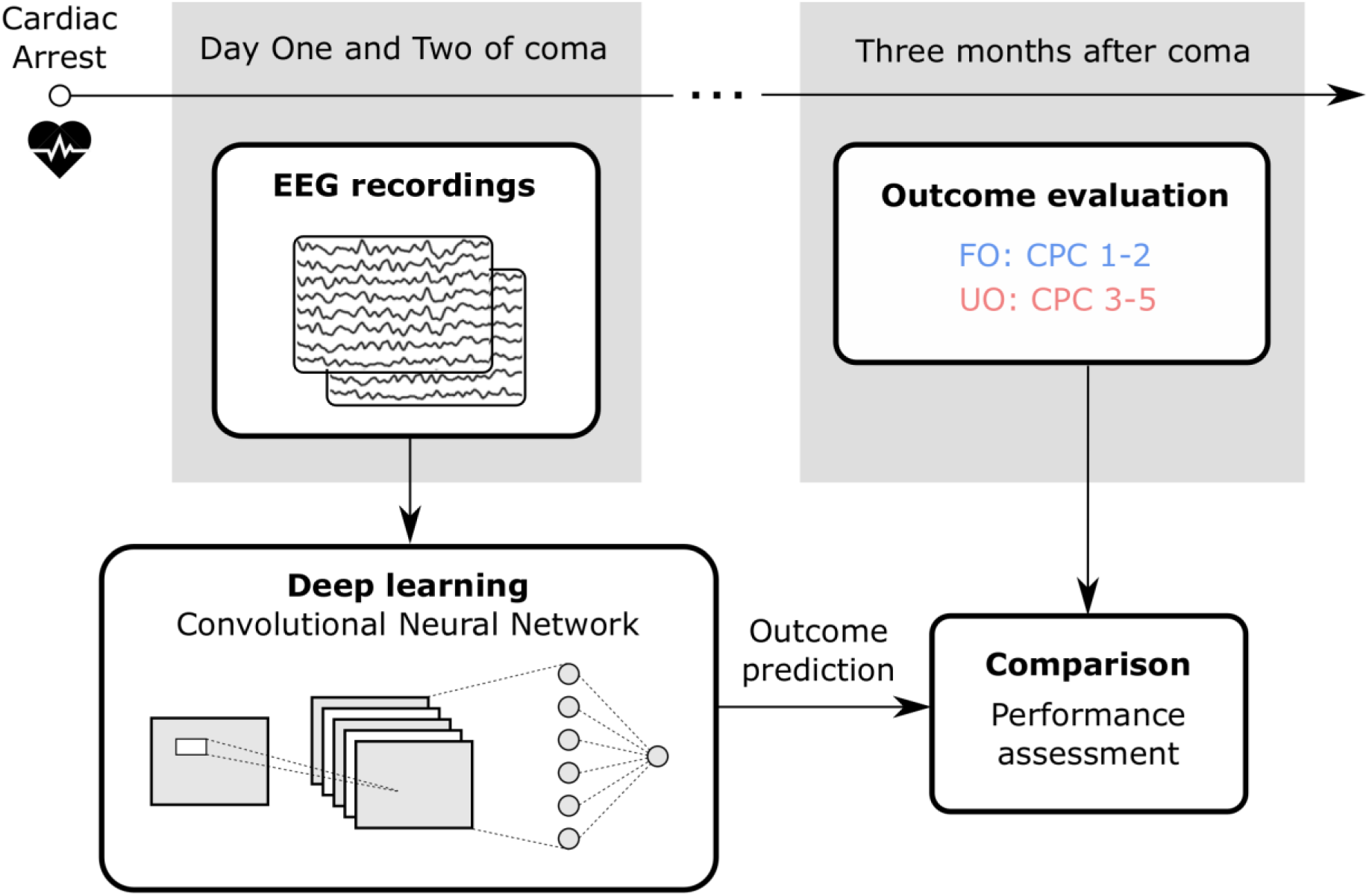

## Introduction

Many patients who are successfully resuscitated from cardiac arrest (CA) do not regain consciousness after return of spontaneous circulation and may remain in a coma for several days or weeks. Early patients’ outcome prognostication plays an important role in the clinical management of comatose patients after CA, since approximately 30% of unfavorable outcome follows withdrawal of life-sustaining therapy in patients with low chances of recovery (Elmer et al., 2016; Leblanc et al., 2018). The current evaluation of a patient’s chances of awakening relies on clinical tests, neurophysiological investigations, brain imaging and serum markers (Edlow et al., 2021), with several tests repeated over the first days of coma. However, these tools mostly focus on predicting unfavorable outcome and for a large number of patients, outcome prognostication remains uncertain early after coma onset (Perkins et al., 2021). Improving early prognosis of patient outcome can lead to personalized treatment, better allocation of intensive care resources and prevent the premature withdrawal of life-sustaining therapy.

The quantitative analysis of resting-state EEG during the first days of coma is emerging as a complementary tool to the existing clinical tests for improving the accuracy and objectivity of patient outcome prediction. A series of studies on resting-state EEG (Beuchat et al., 2018; Kustermann et al., 2019; Pelentritou et al., 2023; Rossetti et al., 2017) and deep learning EEG (Aellen et al., 2023; Pham et al., 2022; Tjepkema-Cloostermans et al., 2019; Zheng et al., 2021) analysis to predict coma patients’ outcome suggested high favorable outcome prediction accuracy in the first 24 hours after coma onset and lower or similar predictive value at later latencies. This evidence suggests that the first day is informative of patient outcome and that later EEG recordings do not substantially improve coma outcome prognostication. Nonetheless, and despite the promise that deep learning analyses offer, coma outcome prediction performance is not yet optimal for clinical application, with area under the receiver operating characteristic (AUC) values ranging from 0.83 and 0.90 (reviewed in (Zubler and Tzovara, 2023)). The aforementioned studies utilized up to 19-channel EEGs, which provide limited spatial information on the neural activity across the scalp compared to high-density EEG systems (i.e., with 62 channels or higher). This leaves unresolved whether, beyond recording latency, deep learning-based coma outcome prognostication could be improved by utilizing a high-resolution EEG.

Here, we evaluated comatose patient outcome prediction on the first and second day after CA using deep learning applied to resting-state EEG. High-density 62-channel EEG recordings were acquired in this study, therefore the performance of our deep learning analysis was evaluated on both the 62-channel and a reduced 19-channel EEG montage, as the latter is more commonly used in the clinical setting. We considered patients with recordings on the first and second day separately, and patients with both recordings, thereby excluding patients who either woke up or died within the first day of coma. Additionally, we assessed the generalizability of our deep learning model using an external dataset acquired in different countries with different patient clinical management and utilizing a variety of EEG systems. Finally, we aimed at evaluating the added value of the deep learning analysis to current clinical predictors of coma outcome and to the EEG power spectrum.

## Methods

The study entitled ‘Mechanisms of post-resuscitation disease after cardiopulmonary arrest in humans’ (23/05; PB_2016-00530) was approved by the local Ethics Committee, *La Commission cantonale d’éthique de la recherche sur l’être humain,* in accordance with the Helsinki Declaration. Informed consent was obtained for all patients in accordance with the ethical approval.

### Patients

Resting-state EEG recordings were collected from comatose patients after CA in Swiss hospitals in Lausanne, Sion, and Bern between 2016 and 2022. EEG data were collected without interruption of clinical management (including sedation) on the first day after coma onset (up to 28 hours after CA) and when possible, on the second day after coma onset (28-56 hours after CA). A second day recording was not performed when: (i) patients awakened; (ii) life support was prematurely withdrawn (following family directive or spontaneous evolution towards brain death); (iii) patients were unavailable due to medical interventions; (iv) experimenters or equipment were unavailable.

During the first day, temperature was controlled using ice packs or cooling pads/cooling catheter with a feedback-controlled cooling device (Arctic Sun System, medivance, Louisville, USA or thermogard XP, ZOLL Medical, Zug, Switzerland). Neuromuscular blocking agents were given to control shivering when needed. Propofol, fentanyl, and/or midazolam were administered for analgesia-sedation (Table 1, Supplementary Table 1). After 24 hours, the temperature was progressively increased to 37°C, muscle relaxants and sedation were weaned off, although a subset of patients was still mildly sedated (Supplementary Table 1). The full outline of unresponsiveness (FOUR (Wijdicks et al., 2005)) score assessed the patients’ functional and neurological state, repeated at least twice within 72 hours.

**Table 1.**
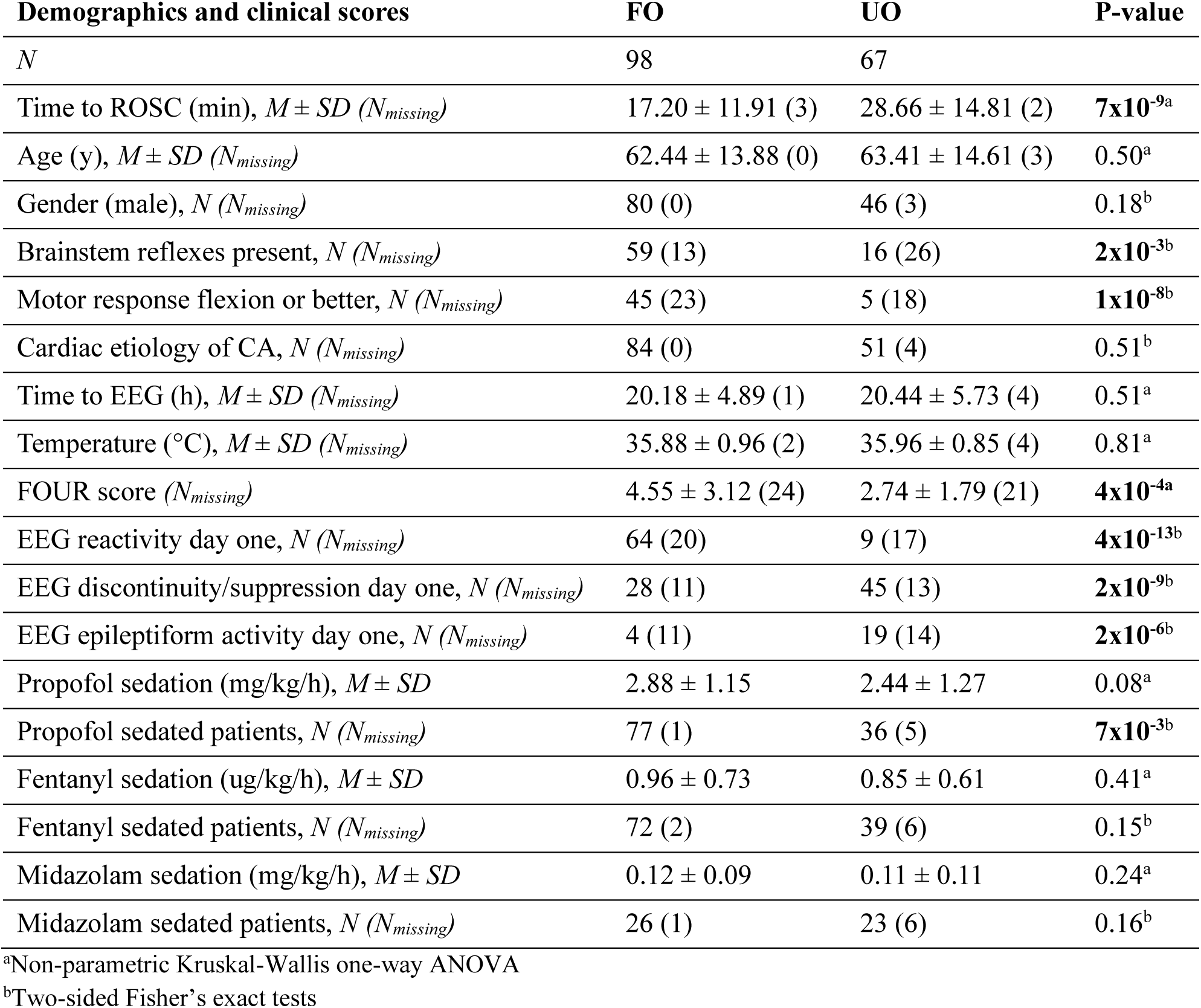
Demographics and clinical characteristics comparison for day one recordings (N = 165)

| Demographics and clinical scores | FO | UO | P-value |
| --- | --- | --- | --- |
| *N* | 98 | 67 |  |
| Time to ROSC (min), $M \pm SD$ ($N_{missing}$) | 17.20 ± 11.91 (3) | 28.66 ± 14.81 (2) | **7x10<sup>-9a</sup>** |
| Age (y), $M \pm SD$ ($N_{missing}$) | 62.44 ± 13.88 (0) | 63.41 ± 14.61 (3) | 0.50<sup>a</sup> |
| Gender (male), *N* ($N_{missing}$) | 80 (0) | 46 (3) | 0.18<sup>b</sup> |
| Brainstem reflexes present, *N* ($N_{missing}$) | 59 (13) | 16 (26) | **2x10<sup>-3b</sup>** |
| Motor response flexion or better, *N* ($N_{missing}$) | 45 (23) | 5 (18) | **1x10<sup>-8b</sup>** |
| Cardiac etiology of CA, *N* ($N_{missing}$) | 84 (0) | 51 (4) | 0.51<sup>b</sup> |
| Time to EEG (h), $M \pm SD$ ($N_{missing}$) | 20.18 ± 4.89 (1) | 20.44 ± 5.73 (4) | 0.51<sup>a</sup> |
| Temperature (°C), $M \pm SD$ ($N_{missing}$) | 35.88 ± 0.96 (2) | 35.96 ± 0.85 (4) | 0.81<sup>a</sup> |
| FOUR score ($N_{missing}$) | 4.55 ± 3.12 (24) | 2.74 ± 1.79 (21) | **4x10<sup>-4a</sup>** |
| EEG reactivity day one, *N* ($N_{missing}$) | 64 (20) | 9 (17) | **4x10<sup>-13b</sup>** |
| EEG discontinuity/suppression day one, *N* ($N_{missing}$) | 28 (11) | 45 (13) | **2x10<sup>-9b</sup>** |
| EEG epileptiform activity day one, *N* ($N_{missing}$) | 4 (11) | 19 (14) | **2x10<sup>-6b</sup>** |
| Propofol sedation (mg/kg/h), $M \pm SD$ | 2.88 ± 1.15 | 2.44 ± 1.27 | 0.08<sup>a</sup> |
| Propofol sedated patients, *N* ($N_{missing}$) | 77 (1) | 36 (5) | **7x10<sup>-3b</sup>** |
| Fentanyl sedation (ug/kg/h), $M \pm SD$ | 0.96 ± 0.73 | 0.85 ± 0.61 | 0.41<sup>a</sup> |
| Fentanyl sedated patients, *N* ($N_{missing}$) | 72 (2) | 39 (6) | 0.15<sup>b</sup> |
| Midazolam sedation (mg/kg/h), $M \pm SD$ | 0.12 ± 0.09 | 0.11 ± 0.11 | 0.24<sup>a</sup> |
| Midazolam sedated patients, *N* ($N_{missing}$) | 26 (1) | 23 (6) | 0.16<sup>b</sup> |

<sup>a</sup>Non-parametric Kruskal-Wallis one-way ANOVA
<sup>b</sup>Two-sided Fisher's exact tests

Pupillary, oculocephalic, corneal reflexes and motor reactivity to pain stimulation were assessed by a certified neurologist at 72 hours. Background reactivity was evaluated by certified readers based on bedside EEG recordings (Tsetsou et al., 2015). Bilateral median nerve somatosensory evoked potentials evaluation was performed 24-28 hours after CA. Further, patients were evaluated using neuron-specific enolase in plasma (NSE), and/or using brain imaging (MRI or CT), depending on the treating physicians’ directive. Withdrawal of life-sustaining therapy was decided using a multimodal approach at least 72 hours after CA, in accordance with the European Society of Intensive Medicine guidelines (Nolan et al., 2021, 2015), when the patient had a Glasgow coma scale motor score of ≤3, and based on at least two of the following: (i) incomplete return of the pupillary and corneal reflexes (Rossetti et al., 2010); (ii) treatment-resistant early myoclonus or status epilepticus (Rossetti et al., 2010); (iii) bilateral absence of cortical somatosensory evoked potentials (Rossetti et al., 2010); (iv) unreactive EEG background activity after sedation weaning (Rossetti et al., 2010); (v) markedly elevated NSE (>60 ug/l) (Rossetti et al., 2010); (vi) extended post-hypoxic lesion in the CT or MRI (Barth et al., 2020). Clinical decisions were independent of and blinded to our retrospective deep learning analysis results.

Patient outcome was defined based on the best functional outcome within three months post CA using the cerebral performance category (CPC (Booth et al., 2004)), a semi-structured phone interview at three months after CA, and the neurological state during hospitalization. Favorable and unfavorable outcomes were assigned to patients scoring a CPC of 1-2 and 3-5, respectively, at any point during the first three months after CA.

### Data acquisition and pre-processing

Continuous resting-state EEG was recorded for 8-20 minutes at 1200 Hz using a 62- active ring electrode array (g.HIamp, g.tec medical engineering, Graz, Austria), referenced to the right ear lobe. EEG data were divided into 5-second segments, down-sampled to 500 Hz and pre-processed using independent component analysis (ICA), artifact rejection, noisy electrode interpolation, and common average re-referencing (Kustermann et al., 2019). The data were finally decimated such that each epoch was separated in five EEG segments of 100 Hz and all values of the initial epoch were only present in one segment, resulting in ‘number of electrodes’ × 100 timepoints as the input to the deep learning analysis for each EEG segment.

### Deep learning using convolutional neural networks

Outcome prediction was evaluated on the first day using the full cohort (N=165), and the first and second day for patients recorded on both days (N=100). In addition, separate models were generated for the 62- channel EEG and a reduced 19-channel montage, the latter being more commonly employed in clinical EEG evaluation. The whole dataset for each day was split in training (64%), validation (16%), and test (20%) with each patient included in only one of the three sets. The result obtained on the test set was representative of a real-life performance since it was not used for training or parameter selection. The consistency of the algorithm performance was evaluated using a five-fold cross-validation procedure with non-overlapping test sets.

A convolutional neural network, ConvNets (O’Shea and Nash, 2015), was used to process the EEG input and generate the coma outcome prediction output. Convolutional neural networks are artificial neural networks that use a set of learnable filters which scan over the input data and extract features at different abstraction levels. These features are then combined to form higher-level representations of the input which are used to classify the input data. The architecture of the ConvNet was inspired by (Schirrmeister et al., 2017), however in our network, the last convolutional layer was replaced by a fully connected layer. This modification allowed for a sufficiently high temporal resolution after the last convolutional layer in order to improve the interpretability of the deep learning results using the GradCAM technique (Selvaraju et al., 2020).

The network was composed of three convolution-max-pooling blocks, wherein the first block was designed to handle the EEG input, followed by two standard blocks. A fully connected layer with 512 neurons was added followed by the last single output neuron. The activation function used in the sub- layers was ReLU (Agarap, 2018). The last layer activation was a sigmoid function giving an output between zero and one which can be interpreted as the predicted probability of favorable outcome. Dropout was used during training in the fully connected layer and set at 0.5 (Srivastava et al., 2014). Batch normalization was used in the convolutional layers, after the activation function (Ioffe and Szegedy, 2015). In total, the model contained 822’350 parameters that were tuned during the training procedure.

The model for each recording day and EEG montage was trained to minimize the binary cross-entropy loss on the train set (Ruby et al., 2020). The optimizer used was Adam with parameters (lr = 5x10^−7^, β = (0.9, 0.999)) and no weight decay (Kingma and Ba, 2015). Mini batches contained 64 EEG segments. The validation set was used to perform early stopping and to select the threshold separating favorable and unfavorable outcome predictions. For early stopping, after each training epoch (one epoch means that each training sample has been used once to update the model parameters), the loss on the validation set was computed and the best validation loss was kept in memory (Prechelt, 1998). Training stopped when the best validation loss did not improve for ten epochs, and the model with the best overall validation loss was retrieved (Prechelt, 1998).

The deep learning analysis produced a prediction result for each five-second EEG segment which was averaged to give the prediction for each subject, recording day, and EEG montage with a threshold aimed at maximizing both favorable and unfavorable prediction. The threshold was the value giving the best geometric mean between the true positive rate (TPR, sensitivity) and the true negative rate (TNR, specificity) on the validation set, as commonly employed for unbalanced datasets (Liu et al., 2013). The threshold was defined as:

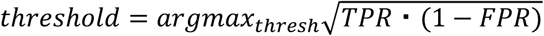

where the TPR and the false positive rate (FPR = 1-TNR) were computed using thresh as threshold. TPR and FPR correspond to the coordinates of a dot on the receiver operating characteristic (ROC) curve on the validation set. The test set was used to compute the performance of the trained network on unseen data.

Performance of the deep learning analysis was evaluated based on the true positive (TP), true negative (TN), false positive (FP) and false negative (FN) outcome predictions where a TP was a favorable outcome patient correctly predicted. The following six metrics were used: AUC, accuracy, sensitivity, specificity, positive predictive value (PPV), and negative predictive value (NPV). Of note, accuracy in this study was defined as the proportion of patients with:

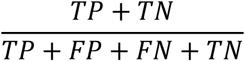

We contrasted the performance on day one and day two as well as for 62-channel and 19-channel models using unpaired t-tests (p<0.05) where the dependent variables were AUC, accuracy, sensitivity, specificity, PPV and NPV across cross-validation folds.

Finally, we used the GradCAM technique to demonstrate which time-points in the EEG signal were identified as features in favor of a favorable or unfavorable outcome prediction (Jonas et al., 2019; Selvaraju et al., 2020). 14 data points were used, representing 14 distinct temporal segments of the EEG segment, each lasting 357 milliseconds.

For all upcoming analyses, a representative cross-validation fold model for each recording day was used to assess whether the performance of our deep learning analysis generalized to data acquired outside Switzerland as well as the complementarity of our results to commonly utilized clinical predictors of comatose patient outcome.

### External test

The performance of the deep learning analysis on the first day of coma was evaluated on an external test set with resting-state EEGs acquired in the Netherlands, Belgium, and United States (Amorim et al., 2023b, 2023a). Since the external dataset included 19-channel EEGs, the first day 19-channel model generated using the Swiss dataset was used to test the performance on the unseen external test dataset. The selected external patients were matched to the Swiss validation and test datasets in the number of patients (N=60), outcome, sex, age, EEG recording latency and number of epochs. External EEG data pre-processing was the same as the Swiss data with the exception of ICA, due to its poor performance with low-density EEG (Cohen, 2014).

### Relationship to other EEG features and clinical markers

The relationship between the deep learning outcome prediction and the 62-channel EEG power spectrum was evaluated using Spearman correlations (p<0.05) between the deep learning predicted probability of favorable outcome for each patient and recording day and the normalized power spectra values between 4.6-15.2 Hz, shown to be predictive of favorable outcome on the first day (Pelentritou et al., 2023).

The relationship between the 62-channel deep learning performance and clinical tests was compared using Mann–Whitney U-tests with Bonferroni correction (p<0.01) between subgroups of patients providing different scores based on the clinical features. The following five binary features were used (Westhall et al., 2016): (i) presence of brainstem reflexes (pupil and corneal reflexes); (ii) presence of motor response (motor Glasgow Coma Scale score ≥4); (iii) reactive EEG background activity; (iv) discontinuous or suppressed EEG background activity (>10% suppression); (v) epileptiform EEG activity (presence of electrographic seizures or status epilepticus, sporadic epileptiform discharges, spiky or sharp periodic discharges or rhythmic spike waves).

Finally, we assessed the deep learning-based outcome prediction for patients with uncertain prediction, namely those for which either none or only one of the clinical evaluators predicted unfavorable outcome.

### Relationship between day one and day two deep learning outcome prediction and sedation

To investigate whether the reduced predictive performance of the 62-channel EEG-based deep learning analysis between the first and second day after coma onset could be explained by differences in the sedative administration between the two days, we computed linear mixed-effects models using the *fitlme* function, as implemented in MATLAB (https://ch.mathworks.com/help/stats/fitlme.html). We investigated the impact of sedation on resting-state EEG in patients with recordings on both days (N=100). To that effect, we generated the following model with predicted probability of favorable outcome as the dependent variable, Propofol Concentration (in mg/kg/h), Midazolam Concentration (in mg/kg/h), Fentanyl Concentration (in µg/kg/h) as fixed factors and Subject as the random factor:

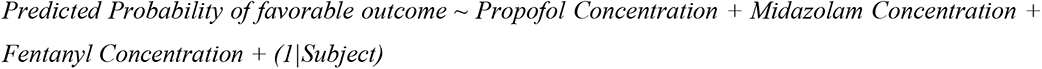

## Results

### Clinical outcome of coma patients

165 patients (outcome: favorable=98, unfavorable=67) were recorded on the first day after coma onset following CA (Table 1) in the University Hospitals of Lausanne (N=67), Sion (N=4), and Bern (N=94). On the second day, we recorded 100 patients (outcome: favorable=51, unfavorable=49) (Supplementary Table 1) in Lausanne (N=46), Sion (N=2) and Bern (N=52).

### Deep learning coma outcome prediction from resting-state EEG

We evaluated the performance of our deep learning models on the first and second day after coma onset and for the 62-channel and 19-channel EEG montage. For the 62-channel model, the average AUC across the five cross-validation folds for the first day of coma (N=165) was 0.93±0.03 (Mean±Standard error) on the test set (Table 2, Figure 1A, Supplementary Figure 1). The accuracy was 0.94±0.03, the PPV 0.93±0.03 and the NPV 0.97±0.02. Similar results were obtained for the first day on the subset of patients recorded on both days (N=100) with an average AUC score of 0.83±0.03 (Supplementary Table 2). Lower predictive performance was observed on the test set for the second day (N=100) compared to the first day for AUC (p=0.04), with a value 0.72±0.05, accuracy (p=0.03), with a value of 0.72±0.05, and PPV (p=0.02), with a value of 0.89±0.04 (Table 2, Figure 1B, Supplementary Figure 1). NPV was not statistically different between the two days (p=0.16) but was still lower than the first day with a value of 0.80±0.09.

**Figure 1.**
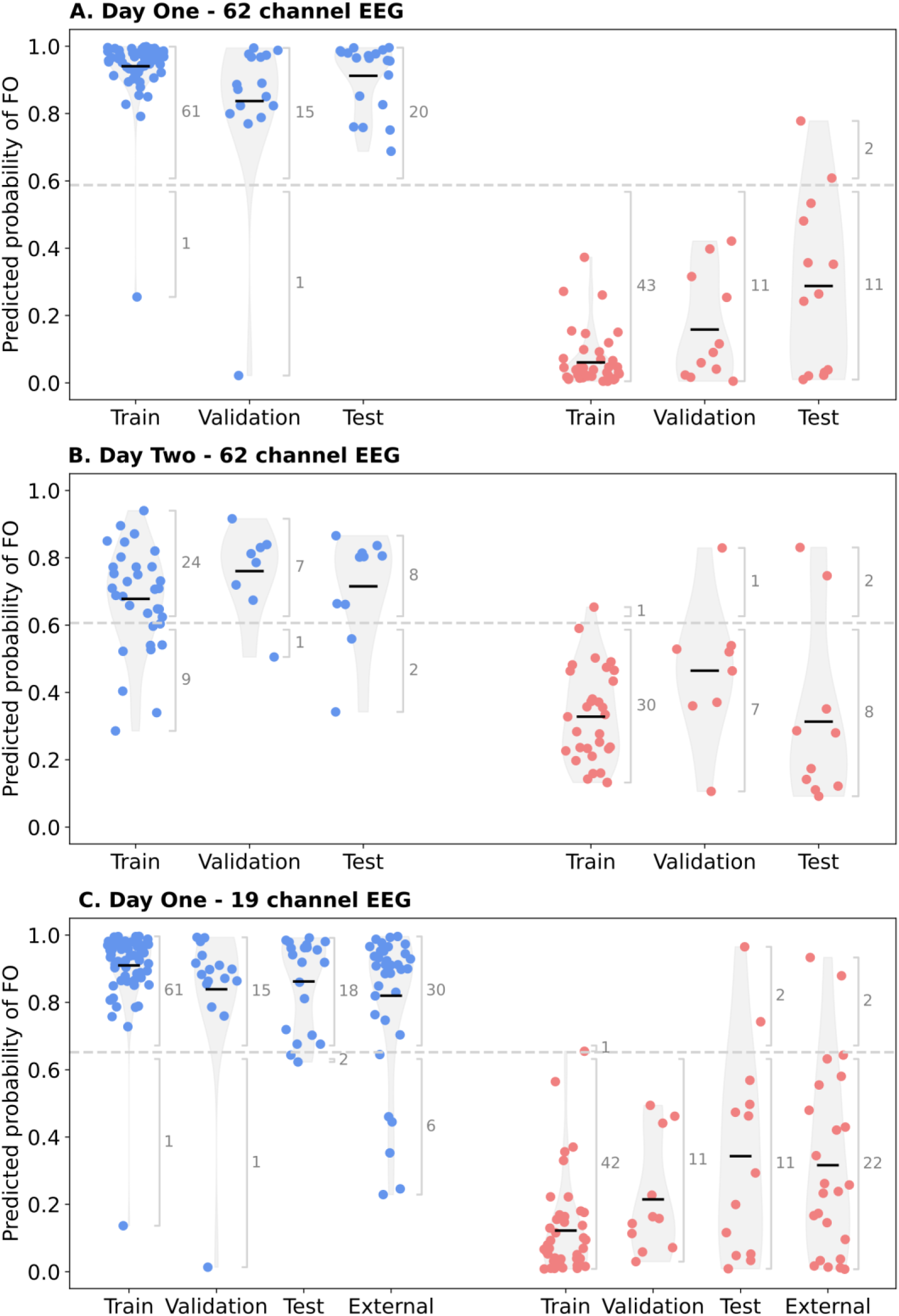
Results for the deep learning analysis applied to comatose patient 62-channel EEG resting- state data (A) from day one (N=165); (B) from day two (N=100); and (C) for 19-channel EEG resting- state data (N=165) and external test data (N=33) on the first day of coma after cardiac arrest. Blue dots correspond to favorable outcome (FO) and red dots to unfavorable outcome (UO) patients. The violin plots (grey areas) show the probability density of the data, smoothed by a kernel density estimator. Black horizontal lines correspond to the means within each plotted population. The results show a high similarity between the validation and test sets suggesting a high degree of model generalization to unseen data. Performance for day one was higher compared to that of day two. The external test results show a high similarity between the Swiss test (Test) and external test (External) sets demonstrating that the deep learning model generalizes to unseen data independent of the acquisition site and equipment.

**Table 2.** Average performance across cross-validation folds for the deep learning analysis applied to the comatose patient 62-channel and 19-channel EEG resting-state data from day one (N=165) and day two (N=100)

| Dataset | AUC<sup>a</sup> | Accuracy<sup>a</sup> | Sensitivity<sup>a</sup> | Specificity<sup>a</sup> | PPV<sup>a</sup> | NPV<sup>a</sup> |
| --- | --- | --- | --- | --- | --- | --- |
| **Day One – 62 channel EEG** |  |  |  |  |  |  |
| Train | $0.97\pm0.01$ | $0.97\pm0.00$ | $0.98\pm0.00$ | $0.97\pm0.01$ | $0.98\pm0.01$ | $0.97\pm0.01$ |
| Validation | $0.94\pm0.02$ | $0.94\pm0.01$ | $0.96\pm0.03$ | $0.91\pm0.05$ | $0.94\pm0.03$ | $0.95\pm0.03$ |
| Test | $0.93\pm0.03$ | $0.94\pm0.03$ | $0.98\pm0.01$ | $0.88\pm0.05$ | $0.93\pm0.03$ | $0.97\pm0.02$ |
| **Day Two – 62 channel EEG** |  |  |  |  |  |  |
| Train | $0.86\pm0.03$ | $0.86\pm0.03$ | $0.82\pm0.06$ | $0.91\pm0.03$ | $0.91\pm0.02$ | $0.84\pm0.05$ |
| Validation | $0.83\pm0.02$ | $0.83\pm0.02$ | $0.88\pm0.04$ | $0.78\pm0.07$ | $0.82\pm0.05$ | $0.88\pm0.03$ |
| Test | $0.72\pm0.05$ | $0.72\pm0.05$ | $0.80\pm0.09$ | $0.63\pm0.04$ | $0.89\pm0.04$ | $0.80\pm0.09$ |
| **Day One – 19 channel EEG** |  |  |  |  |  |  |
| Train | $0.98\pm0.00$ | $0.98\pm0.00$ | $0.98\pm0.00$ | $0.98\pm0.01$ | $0.99\pm0.01$ | $0.97\pm0.01$ |
| Validation | $0.93\pm0.02$ | $0.93\pm0.02$ | $0.95\pm0.02$ | $0.91\pm0.05$ | $0.94\pm0.03$ | $0.93\pm0.03$ |
| Test | $0.89\pm0.03$ | $0.90\pm0.03$ | $0.95\pm0.02$ | $0.84\pm0.05$ | $0.90\pm0.03$ | $0.92\pm0.03$ |
| **Day Two – 19 channel EEG** |  |  |  |  |  |  |
| Train | $0.91\pm0.03$ | $0.91\pm0.03$ | $0.86\pm0.07$ | $0.96\pm0.04$ | $0.97\pm0.03$ | $0.88\pm0.05$ |
| Validation | $0.85\pm0.03$ | $0.85\pm0.03$ | $0.88\pm0.00$ | $0.83\pm0.06$ | $0.85\pm0.05$ | $0.87\pm0.01$ |
| Test | $0.75\pm0.03$ | $0.76\pm0.03$ | $0.78\pm0.11$ | $0.73\pm0.08$ | $0.77\pm0.02$ | $0.83\pm0.08$ |

<sup>a</sup>Expressed as Mean ± Standard error

For the test set, we obtained a lower performance for the 19-channel model compared to the 62-channel model on the first day (N=165) for AUC (p=5.63x10^-4^) with a value of 0.89±0.03, accuracy (p=3.90x10^-3^), with a value of 0.90±0.03, and PPV (p=0.04), with a value of 0.90±0.03 (Table 2, Figure 1C). Despite a lower NPV value of 0.92±0.03 for the 19-channel model, this was not significantly different to the 62- channel model (p=0.20). No differences (p>0.05) were observed between the 62-channel and 19-channel models on the second day (Table 2).

Our deep learning analysis was further evaluated on a subset of patients (N=60) from an external 19- channel EEG dataset on the first day of coma (Amorim et al., 2023b, 2023a). Considering the better performance on day one, our first day 19-channel model for a representative cross-validation fold was tested using the external dataset (Table 3). We obtained an outcome prediction with high performance on the Swiss and external test sets with an AUC of 0.87 and 0.88, respectively and accuracy of 0.88 and 0.87, respectively (Table 3, Figure 1C). PPV and NPV were also comparable, with PPV of 0.90 and 0.94 and NPV of 0.85 and 0.79 for the Swiss and external test sets, respectively (Table 3).

**Table 3.** Performance of the deep learning analysis for a representative cross-validation fold applied to the comatose patient EEG resting-state data with 62-channels on day one (N=165) and day two (N=100), with 62-channels for the subset of patients with inconsistent clinical markers for unfavorable outcome prediction on day one (N=53) and day two (N=43), and with 19-channels on day one for the Swiss (N=165) and external data (N=60)

| Dataset | TP/FP/FN/TN | AUC | Accuracy | Sensitivity | Specificity | PPV | NPV |
| --- | --- | --- | --- | --- | --- | --- | --- |
| **Day One – 62 channel EEG** |  |  |  |  |  |  |  |
| Train | 61/0/1/43 | 0.99 | 0.99 | 0.98 | 1.00 | 1.00 | 0.98 |
| Validation | 15/0/1/11 | 0.97 | 0.96 | 0.94 | 1.00 | 1.00 | 0.92 |
| Test | 20/2/0/11 | 0.92 | 0.94 | 1.00 | 0.85 | 0.91 | 1.00 |
| Uncertain outcome | 50/0/1/2 |  | 0.98 | 0.98 | 1.00 | 1.00 | 0.67 |
| **Day Two – 62 channel EEG** |  |  |  |  |  |  |  |
| Train | 24/1/9/30 | 0.85 | 0.84 | 0.73 | 0.97 | 0.96 | 0.77 |
| Validation | 7/1/1/7 | 0.88 | 0.88 | 0.88 | 0.88 | 0.88 | 0.88 |
| Test | 8/2/2/8 | 0.80 | 0.80 | 0.80 | 0.80 | 0.80 | 0.80 |
| Uncertain outcome | 26/2/9/6 |  | 0.74 | 0.74 | 0.75 | 0.93 | 0.40 |
| **Day One – 19 channel EEG** |  |  |  |  |  |  |  |
| Swiss Train | 61/1/1/42 | 0.98 | 0.98 | 0.98 | 0.98 | 0.98 | 0.98 |
| Swiss Validation | 15/0/1/11 | 0.97 | 0.96 | 0.94 | 1.00 | 1.00 | 0.92 |
| Swiss Test | 18/2/2/11 | 0.87 | 0.88 | 0.90 | 0.84 | 0.90 | 0.85 |
| External Test | 30/2/6/22 | 0.88 | 0.87 | 0.83 | 0.92 | 0.94 | 0.79 |

### Deep learning-based outcome prediction, EEG features, and clinical markers

The predictive performance of the deep learning analysis was compared to that of 62-channel resting-state EEG normalized power spectra between 4.6–15.2 Hz (Pelentritou et al., 2023). The predicted probability of favorable outcome and the EEG power spectra (Figure 2) positively correlated for favorable (r=0.34, p=0.01) and negatively correlated for unfavorable (r=-0.28, p=0.02) outcome on day one. No correlation was observed for a prediction of favorable (r=-0.14, p=0.33) or unfavorable (r=0.02, p=0.88) outcome on day two.

**Figure 2.**
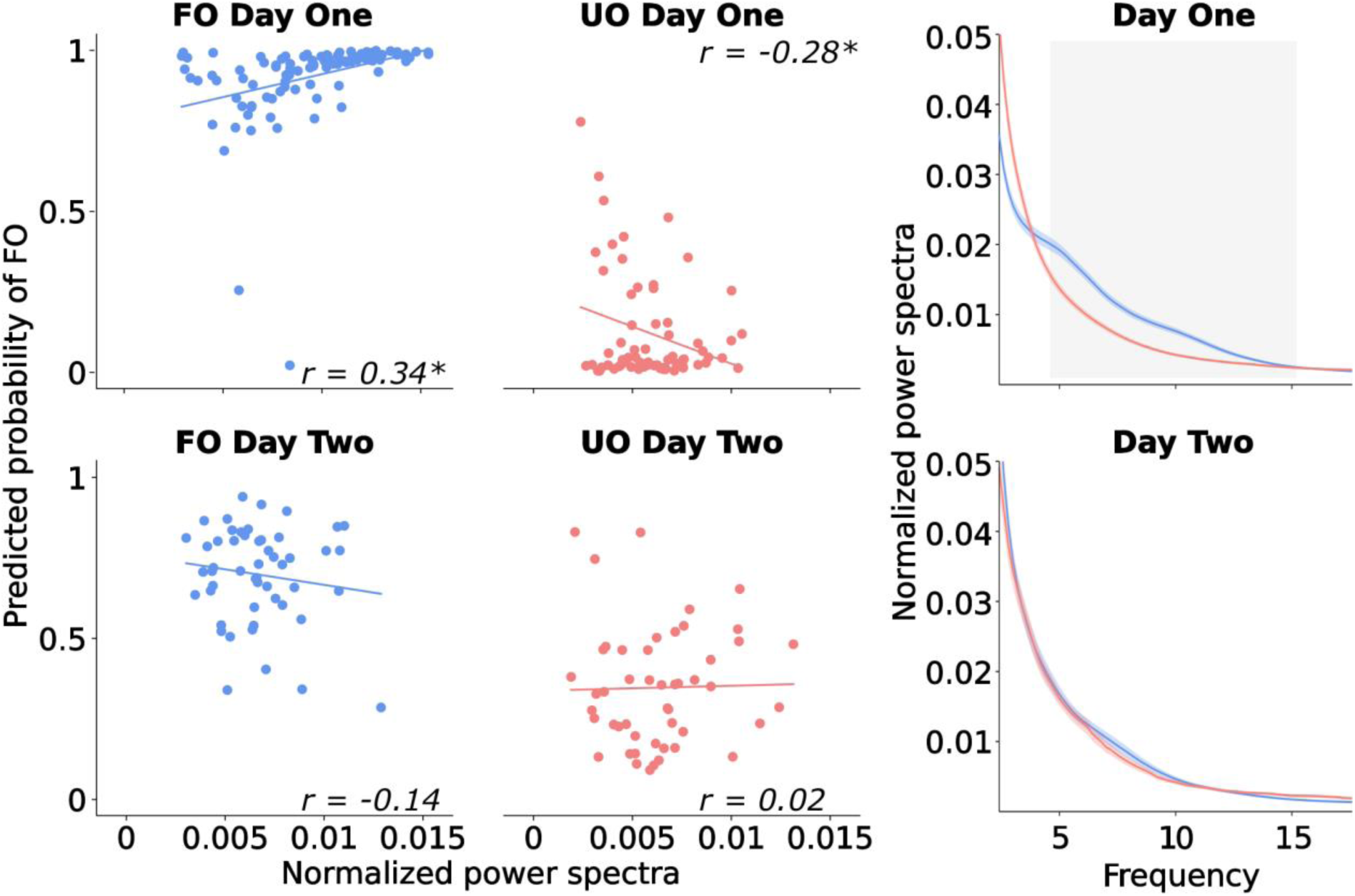
Correlation between the predicted probability of favorable outcome (FO) based on the deep learning analysis and normalized spectral power in the 4.6-15.2 Hz range using 62-channel EEG acquired on the first (N=165) and second day (N=100) of coma for FO and unfavorable outcome (UO) patients. Normalized power spectra were computed for each recording day by averaging the power spectrum between 2 and 40 Hz across artifact-free 5-second segments and normalizing based on the sum of all spectral values. The normalized power spectra between 4.6 Hz and 15.2 Hz were then extracted and are presented in the figure in arbitrary units. Blue dots correspond to FO and red dots to UO patients. Spearman correlation r values between predicted probability of FO and the normalized power spectra are shown for each day and patient population with any significant correlations marked with an asterisk (p<0.05, *). The corresponding group averaged power spectrum for FO and UO patients on the first and second day of coma are plotted with shaded areas indicating the standard error of the mean across patients. FO patients on the first day of coma exhibit a significant correlation with the model’s prediction, suggesting that for this subgroup of patients, the deep learning analysis utilizes EEG features related to the power spectra in predicting the patient outcome.

We contrasted the 62-channel deep learning performance on day one to commonly utilized clinical features for coma outcome prediction (Figure 3). The deep learning predicted probability of favorable outcome on day one between patients with favorable and unfavorable clinical features differed in reactivity (p=1.98x10^-10^), discontinuity (p=3.16x10^-9^), epileptiform features (p=1.01x10^-5^) and motor response (p=8.30x10^-7^), but not brainstem reflexes (p=2.11x10^-2^). The correspondence between the deep learning results and clinical features was verified using the GradCam inspection of EEG segments (Figure 4).

**Figure 3.**
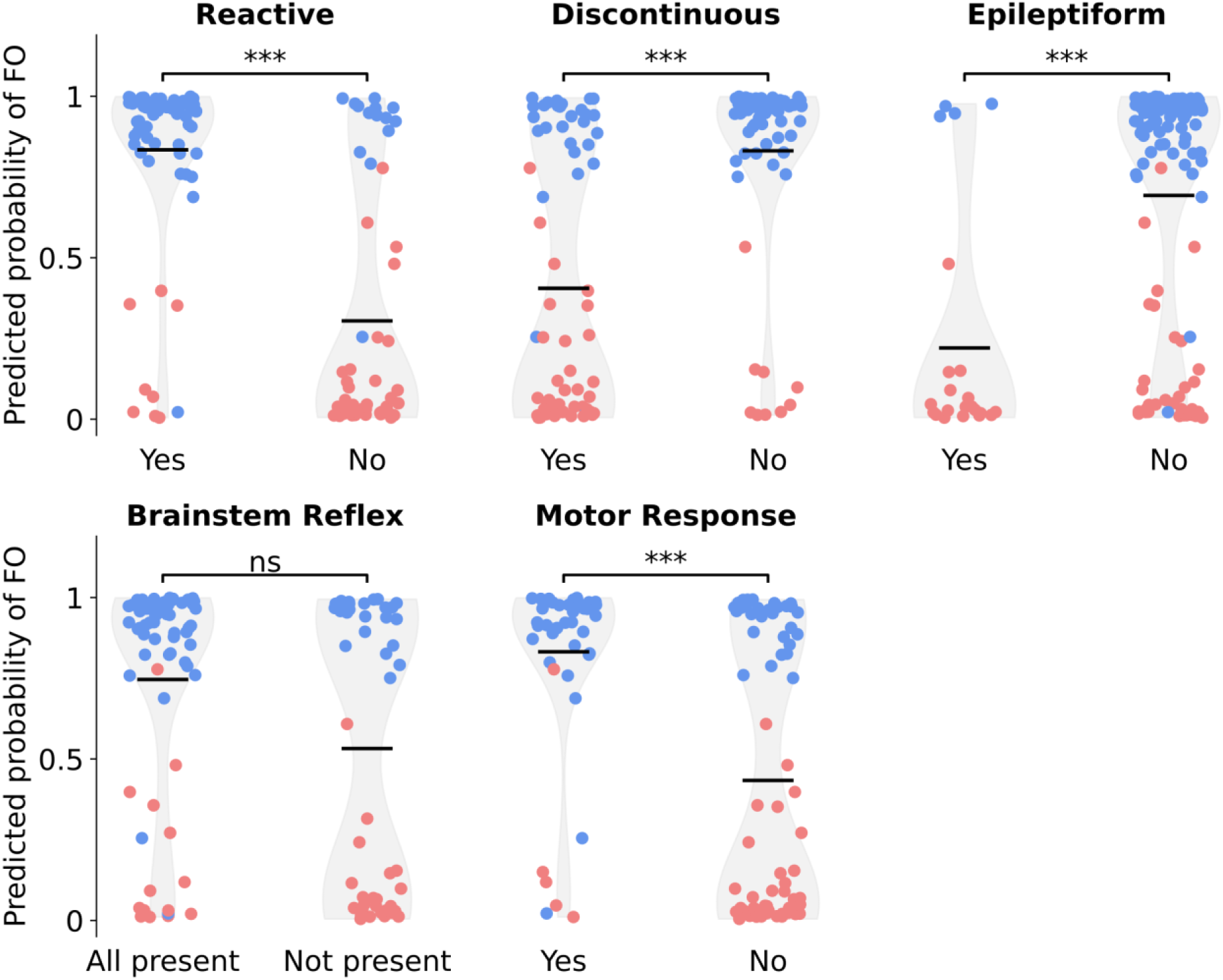
The deep learning analysis output according to the different features used for clinical prognosis on the first day after coma onset. Blue dots correspond to favorable outcome (FO) and red dots to unfavorable outcome (UO) patients. The vertical axis shows the predicted probability of FO by the deep learning analysis calculated using 62-channel resting-state EEG data from the first day after coma onset. The grey shaded area is a violin plot demonstrating the probability density of the data, smoothed by a kernel density estimator. Black horizontal bars show the mean predicted probability of FO across the plotted population. Model performance was statistically compared using Mann–Whitney U-tests with Bonferroni correction for patients with and without: Reactive EEG (p<0.0001), Discontinuous EEG (p<0.0001), Epileptiform EEG features (p<0.0001), Brainstem Reflexes (not significant (ns)), Motor response (p<0.0001). A significant difference between clinical features was observed in all cases except for Brainstem Reflexes.

**Figure 4.**
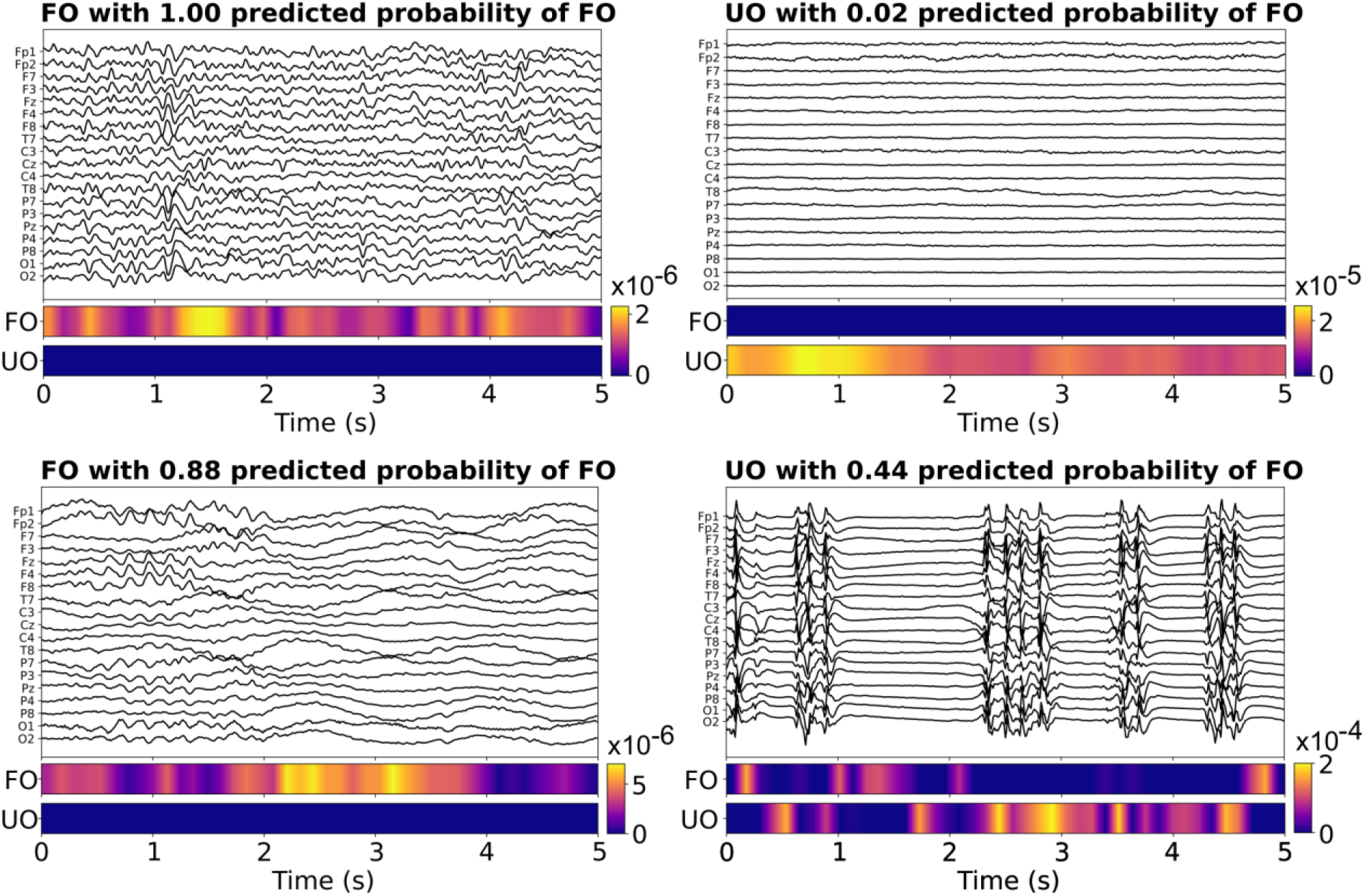
EEG features identified by the deep learning analysis in classifying favorable and unfavorable outcome. Exemplar five-second EEG segments for the reduced 19-electrode montage (for better visualization) from four correctly classified patients with favorable outcome (FO) and unfavorable outcome (UO). The heatmaps indicate which temporal segments are discriminative of FO or UO as recognized by the Grad-CAM algorithm. The color bar corresponds to the degree of discriminative significance a region holds for a specific patient outcome. The top left panel shows a continuous EEG pattern with well-modulated alpha (8-12 Hz) activity which results in a perfect FO prediction of 1.0. The bottom left panel demonstrates generalized rhythmic theta (∼6 Hz) followed by delta (<1 Hz) rhythmic activity, both discriminative of FO. The top right panel demonstrates a suppressed EEG with no distinct electrophysiological features in a UO patient, which yields a near perfect prediction of UO. The bottom right panel shows burst suppression with highly epileptiform bursts which are largely indicative of UO, as correctly identified by the neural network; of note, slow deflections in the transition between suppressed and burst parts could be suggestive of FO for the convolutional neural network. The four panels demonstrate the correspondence between the classic EEG features that are indicative of coma outcome based on clinical visual inspection, also considered by the deep learning analysis.

Finally, we showed that the deep learning analysis of the 62-channel resting-state EEG is informative in patients with uncertain outcome on the first day. We defined this subset of patients as those who had none or one of five clinical features of unfavorable outcome (Figure 3). We identified 53 (32%) uncertain outcome patients on day one and 43 (43%) on day two. In this subset, accuracy for correct patient classification was higher on day one at 0.98 compared to day two at 0.74 (Table 3).

### Deep learning-based outcome prediction and sedation

We assessed the relationship between the predictive performance of the deep learning analysis and the level of sedation (which could impact the resting-state EEG features) on day one and day two using a linear mixed model. This analysis revealed no significant main effects of Propofol Concentration (p=0.07), Midazolam Concentration (p=0.24), and Fentanyl Concentration (p=0.14) on the predicted probability of favorable outcome.

## Discussion

We investigated the prognostic performance of a deep learning analysis of the 62-channel EEG in comatose patients after CA during the first two days of coma, collected across three Swiss hospitals. The best predictive performance was observed using data from the first day with an average accuracy of 0.94±0.03 across cross-validation folds evaluated in a test dataset. The second day provided less accurate prediction with an average accuracy of 0.72±0.05. Prediction performance was higher with the 62-channel compared to the downsampled 19-channel EEG which produced an average accuracy of 0.90±0.03, suggesting that spatial information relevant for the EEG-based prediction of coma outcome might be hindered by a low-density montage. Testing the 19-channel model on a dataset acquired outside Switzerland validated our algorithm performance on the first day of coma with a high accuracy of 0.87. High prediction accuracy for the 62-channel model extended to patients with uncertain outcome, with an accuracy of 0.98 on the first day of coma. Finally, the first day deep learning-based prognosis correlated with the 62-channel EEG power spectrum (4.6-15.2 Hz) and provided complementary information to the patient clinical evaluation, including markers based on malignant EEG patterns and motor responses.

### Coma outcome prediction using deep learning

Our results add to the literature demonstrating that deep learning analyses of resting-state EEG (Jonas et al., 2019; Pham et al., 2022; Tjepkema-Cloostermans et al., 2019; van Putten et al., 2018; Zheng et al., 2021) may provide higher predictive performance of comatose patient outcome after CA compared to studies that use predefined resting-state EEG features such as power spectra (Kustermann et al., 2019; Pelentritou et al., 2023), functional connectivity (Beudel et al., 2014; Kustermann et al., 2020; Zubler et al., 2017), or malignant patterns (Müller et al., 2020). Previous reports of deep learning analyses applied to resting-state EEG reported lower AUC values compared to our obtained value of 0.93±0.03: an AUC of 0.89 on the first day of coma (Jonas et al., 2019), an AUC of 0.88 at 24 hours after coma onset (Tjepkema-Cloostermans et al., 2019), the best AUC of 0.89 at 24 hours after coma onset (Zheng et al., 2021) and an AUC between 0.88-0.90 within the first day after coma onset (Pham et al., 2022). The lower performance in these studies could be explained by differences in the deep learning algorithm implementation. These include: (i) when training was stopped and the smaller number of deep learning parameters used to optimize the deep learning algorithm (Jonas et al., 2019); (ii) the type of algorithm used (Tjepkema-Cloostermans et al., 2019; Zheng et al., 2021); (iii) the data used to train the model, herein we trained separately for day one and day two, differently from (Pham et al., 2022). Importantly, our results demonstrate that a higher density EEG montage of 62 channels serving as input for the deep learning analysis can also improve algorithm performance, since the lower density 19-channel montage produced a lesser AUC of 0.89±0.03, closer to values obtained in previous studies.

In agreement with a growing literature demonstrating a higher predictive performance of EEG-based markers on the first compared to the second day of coma (Pelentritou et al., 2023; Tjepkema-Cloostermans et al., 2019), we obtained a better prognostication on the first day. This decrease from the first to the second day can be explained by the heterogeneity in the patterns of recovery for favorable outcome patients and of neural degeneration in unfavorable outcome patients. Additional factors that could explain the difference in prediction accuracy between the two days relate to the patients’ clinical management, mainly consisting of differences in sedation (Pelentritou et al., 2023). In our cohort, differences in the level of sedation did not seem to impact the deep learning outcome prediction, nonetheless, the systematic evaluation of such divergent clinical care between the two days is precluded by the current clinical guidelines for patient management.

### Complementarity between deep learning and EEG clinical features

The 62-channel resting-state EEG power spectrum between 4.6-15.2 Hz in favorable and unfavorable outcome patients on the first day of coma correlated with the deep learning scores. These results align with previous results showing the utility of the EEG power spectra in predicting some of the patients with FO on the first day of coma (Kustermann et al., 2019; Pelentritou et al., 2023). Despite reduced predictive performance, the deep learning algorithm was still predictive of outcome on the second day of coma, suggesting that outcome prediction on the second day relies on resting-state EEG features beyond spectral features.

We additionally observed an overlap between the deep learning results and the EEG-based clinical features, namely discontinuity and epileptiform features, suggesting that the algorithm may exploit similar patterns to the clinical assessment. These results are consistent with the prevalence of malignant EEG patterns (e.g., suppressed background, burst-suppression – especially if showing identical bursts -, epileptiform activity) in unfavorable outcome patients and of continuous background activity in favorable outcome patients (Hofmeijer et al., 2015; Westhall et al., 2016). The overlap between the deep learning results and EEG reactivity and motor response is less obvious. These clinical features are not assessed using resting-state EEG; reactivity is evaluated upon noxious stimulation and the motor response does not rely on EEG-based evaluation. This result suggests some interdependency between features of the resting-state EEG and these markers’ assessment. The lack of a relationship between the deep learning analysis and brainstem reflexes resembles earlier findings using a multivariate analysis of EEG responses to sounds which identified patients who could not be predicted upon clinical evaluation, including patients without preserved brainstem reflexes (Tzovara et al., 2016).

### Limitations

A limitation for this work is the smaller sample size in our cohort acquired over years with changing clinical practices compared to previous studies (Jonas et al., 2019; Tjepkema-Cloostermans et al., 2019). In addition, we acknowledge that some clinical features commonly utilized by treating physicians were not collected as part of this work and therefore, could not be evaluated in relation to the deep learning results. These include, but are not limited to, information on brain imaging, pupillometry, and epileptiform features subtypes (Ruijter et al., 2022).

## Conclusions

We obtained high predictive performance of comatose patient outcome during the first day after CA using a deep learning analysis of high-density resting-state EEG data. Outcome prognostication outperformed the results obtained with a lower density EEG montage and the clinical prediction in the difficult cases where outcome was uncertain using clinical markers, yielding similar prediction accuracy to the entire cohort. Importantly, while current clinical evaluation of comatose patient outcome mostly focuses on unfavorable outcome prognostication (Nolan et al., 2021), our results align with recent efforts to improve early favorable outcome prediction (Vanat et al., 2023).

## Supporting information

Supplementary Materia

## Data Availability

The datasets generated during the current study are not publicly available due to the sensitive nature of the clinical data.

## Acknowledgements

We thank all EEG technicians from the involved hospitals for invaluable help with EEG recordings. The authors are thankful to Nathalie Ata Nguissi and Thomas Kustermann for data collection, data pre- processing, and inspiring previous work. We are indebted to Rupert Ortner and Christoph Guger for technical support, and to Laura Pezzi and Yoanne Boulez for their help in data acquisition.

## Author contributions

**Andria Pelentritou:** Conceptualization; Data curation; Methodology; Formal analysis; Writing - original draft, **Lucas Gruaz:** Conceptualization; Data curation; Methodology; Formal analysis; Writing - original draft, **Manuela Iten:** Resources; Visualization; Writing - review & editing, **Matthias Haengii:** Resources; Visualization; Writing - review & editing, **Frederic Zubler:** Resources; Visualization; Writing - review & editing, **Andrea O Rossetti:** Resources; Visualization; Writing - review & editing, **Marzia De Lucia:** Conceptualization; Methodology; Funding acquisition; Project administration; Supervision; Writing - original draft.

