## Supplementary Materia for "High density EEG and deep learning improves outcome prediction on the first day of coma after cardiac arrest"

MP16 05 559, Chemin de Mont-Paisible 16, 1011 Lausanne, Switzerland

### Supplementary Tables

**Supplementary Table 1. Demographics and clinical characteristics of favorable outcome (FO) and unfavorable outcome (UO) patients with day one and day two recordings (N = 100)**

| | **FO** | **UO** | **P-value** |
|---|---|---|---|
| *N* | 51 | 49 | |
| Time to ROSC (min), $M \pm SD$ ($N_{missing}$) | 17.52 ± 12.54 (0) | 28.39 ± 13.48 (0) | **$8x10^{-6}$**[a] |
| Age (y), $M \pm SD$ ($N_{missing}$) | 63.04 ± 12.99 (0) | 66.06 ± 12.44 (0) | 0.18[a] |
| Gender (male), $N$ ($N_{missing}$) | 43 (0) | 35 (0) | 0.15[b] |
| Brainstem reflexes present, $N$ ($N_{missing}$) | 39 (2) | 13 (17) | **$7x10^{-4}$**[b] |
| Motor response flexion or better, $N$ ($N_{missing}$) | 29 (3) | 5 (8) | **$3x10^{-6}$**[b] |
| Cardiac etiology, $N$ ($N_{missing}$) | 40 (0) | 36 (3) | 1.00[b] |
| Time to EEG day one (h), $M \pm SD$ ($N_{missing}$) | 19.28 ± 4.67 (0) | 20.24 ± 5.42 (0) | 0.23[a] |
| Temperature day one (°C), $M \pm SD$ ($N_{missing}$) | 35.82 ± 1.03 (0) | 35.95 ± 0.79 (1) | 0.98[a] |
| FOUR score day one ($N_{missing}$) | 4.16 ± 2.53 (6) | 2.75 ± 1.81 (13) | **$5x10^{-3}$**[a] |
| EEG reactivity day one, $N$ ($N_{missing}$) | 36 (6) | 5 (13) | **$2x10^{-9}$**[b] |
| EEG discontinuity/suppression day one, $N$ ($N_{missing}$) | 18 (7) | 33 (9) | **$1x10^{-4}$**[b] |
| EEG epileptiform activity day one, $N$ ($N_{missing}$) | 0 (7) | 17 (10) | **$3x10^{-7}$**[b] |
| Propofol sedation day one (mg/kg/h), $M \pm SD$ | 2.58 ± 1.23 | 2.33 ± 1.30 | 0.40[b] |
| Propofol sedated patients day one, $N$ ($N_{missing}$) | 40 (1) | 31 (0) | 0.08 [b] |
| Fentanyl sedation (ug/kg/h) day one, $M \pm SD$ | 0.92 ± 0.61 | 0.80 ± 0.61 | 0.20[a] |
| Fentanyl sedated patients day one, $N$ ($N_{missing}$) | 43 (1) | 31 (0) | **0.01**[b] |
| Midazolam sedation (mg/kg/h) day one, $M \pm SD$ | 0.13 ± 0.10 | 0.12 ± 0.12 | 0.35[a] |
| Midazolam sedated patients day one, $N$ ($N_{missing}$) | 20 (1) | 18 (0) | 0.84[b] |
| Time to EEG day two (h), $M \pm SD$ ($N_{missing}$) | 42.96 ± 5.03 (0) | 44.08 ± 5.73 (0) | 0.16[a] |
| Temperature day two (°C), $M \pm SD$ ($N_{missing}$) | 36.98 ± 0.86 (0) | 36.94 ± 0.68 (1) | 0.45[a] |
| FOUR score day two ($N_{missing}$) | 6.93 ± 3.84 (9) | 3.80 ± 2.84 (10) | **$3x10^{-5}$**[a] |
| EEG reactivity day two, $N$ ($N_{missing}$) | 39 (8) | 12 (12) | **$8x10^{-8}$**[b] |
| EEG discontinuity/suppression day two, $N$ ($N_{missing}$) | 3 (6) | 16 (9) | **$5x10^{-4}$**[b] |
| EEG epileptiform activity day two, $N$ ($N_{missing}$) | 2 (6) | 11 (12) | **$2x10^{-3}$**[b] |
| Propofol sedation day two (mg/kg/h), $M \pm SD$ | 2.31 ± 1.42 | 2.03 ± 1.40 | 0.56[a] |
| Propofol sedated patients day two, $N$ ($N_{missing}$) | 30 (3) | 19 (0) | **0.03**[b] |
| Fentanyl sedation (ug/kg/h) day two, $M \pm SD$ | 1.55 ± 1.57 | 0.87 ± 0.77 | 0.11[a] |
| Fentanyl sedated patients day two, $N$ ($N_{missing}$) | 22 (3) | 11 (0) | **0.02**[b] |
| Midazolam sedation (mg/kg/h) day two, $M \pm SD$ | 0.09 ± 0.05 | 0.20 ± 0.24 | 0.62[a] |
| Midazolam sedated patients day two, $N$ ($N_{missing}$) | 4 (3) | 5 (0) | 1.00[b] |

[a]Non-parametric Kruskal-Wallis one-way ANOVA
[b]Two-sided Fisher's exact tests

**Supplementary Table 2. Average performance of the deep learning analysis across cross-validation folds applied to the comatose patient 62-channel EEG resting-state data on day one for patients recorded on both days (N=100)**

| Dataset | AUC[a] | Accuracy[a] | Sensitivity[a] | Specificity[a] | PPV[a] | NPV[a] |
|---|---|---|---|---|---|---|
| **Day One (N=100)** | | | | | | |
| Train | 0.97±0.01 | 0.97±0.01 | 0.98±0.01 | 0.97±0.01 | 0.97±0.02 | 0.97±0.01 |
| Validation | 0.88±0.04 | 0.88±0.04 | 0.88±0.04 | 0.88±0.06 | 0.88±0.05 | 0.88±0.04 |
| Test | 0.83±0.03 | 0.83±0.03 | 0.88±0.04 | 0.77±0.07 | 0.82±0.05 | 0.87±0.04 |

[a]Expressed as Mean ± Standard error

### Supplementary Figures

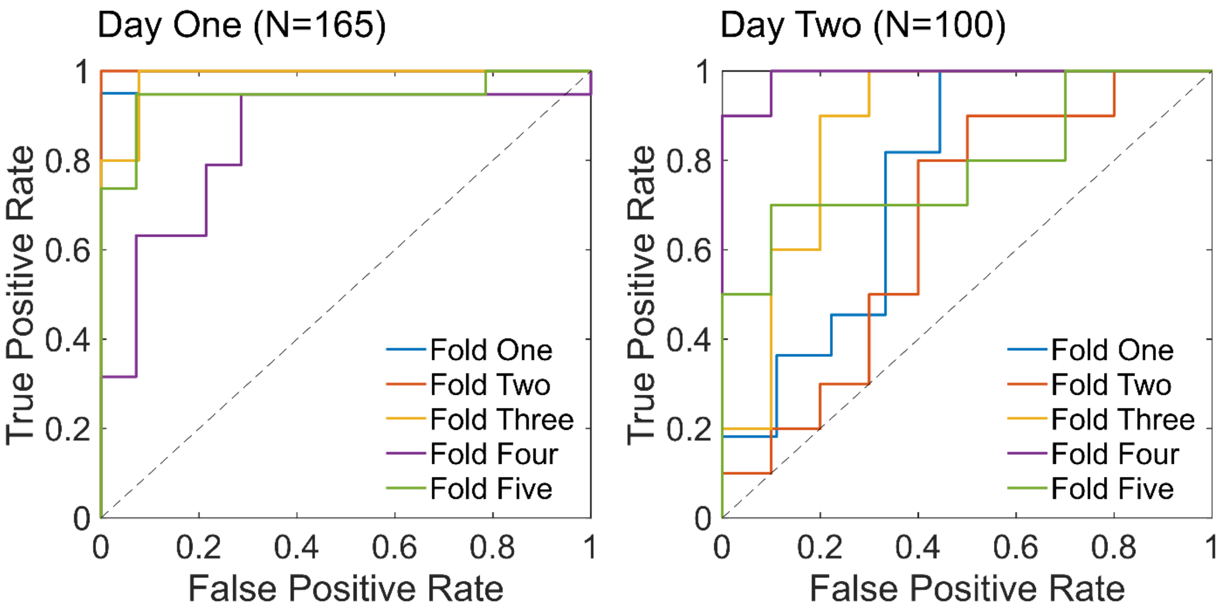


**Supplementary Figure 1. Receiver operating characteristic curves for the deep learning analysis applied to the comatose patient 62-channel EEG resting-state data on day one and day two.** Colored lines represent the predicted True Positive Rate and False Positive Rate in the patients from day one (N=165) and day two (N=100) for the five folds of cross-validation test sets.
